## Supplementary file1 for "Child Support Grant expansion and cognitive function among women in rural South Africa: findings from a natural experiment in HAALSI cohort"

Supplementary Materials

**A. Child Support Grant Variable Construction**

Detailed below are the steps we took to construct the duration of Child Support Grant (CSG) eligibility exposure variable used in the main analysis:

*Step 1: Plotting the relationship between all years of CSG eligibility (indicator variables, range of CSG eligibility years: 1 to 98.5) and cognitive function z-scores*

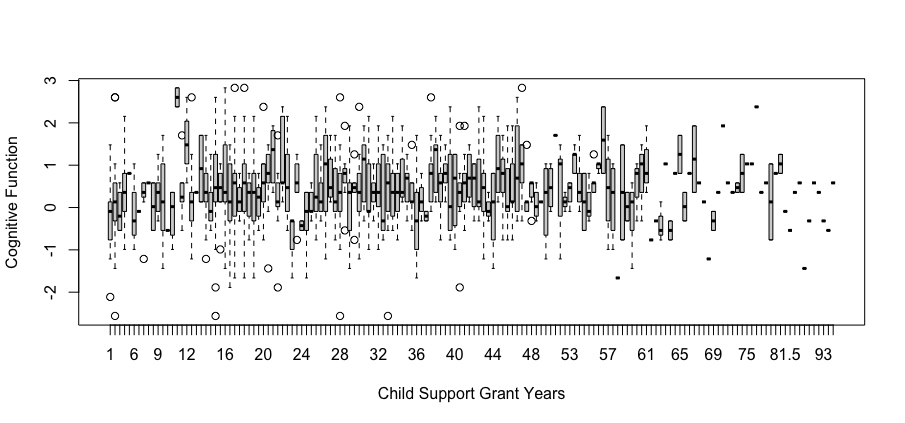
 **S1 Fig: Plotting the relationship between all years of CSG eligibility and cognitive function z-scores**

*Step 2: Plotting the relationship between years of CSG eligibility in five-year intervals and cognitive function z scores [group 1: 1 to 5 years of CSG eligibility; group 2: 5.5 to 10 years, group 3: 10.5 to 15 years, group 4: 15.5 to 20 years and so on]*

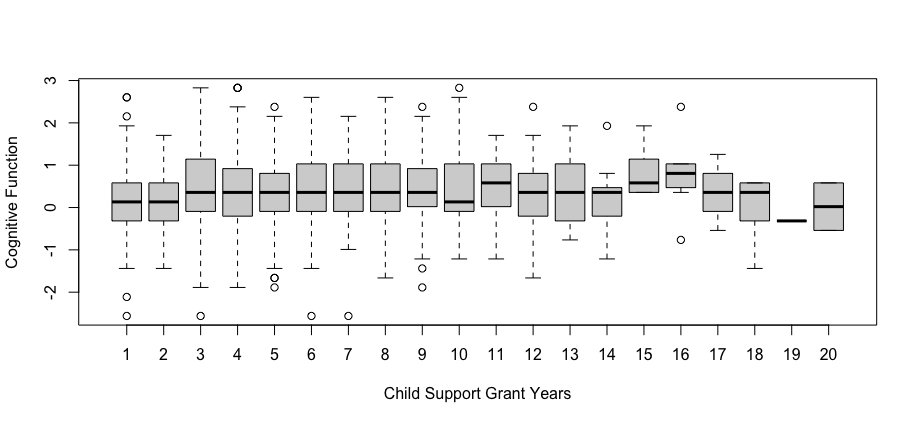

**S2 Fig: Plotting the relationship between years of CSG eligibility in five-year intervals and cognitive function z scores**

*Step 3: Plotting the relationship between years of CSG eligibility in five-year intervals and cognitive function z scores [after collapsing the CSG eligibility groups from 50.5 years and more to one group (group 11) due to small sample size in each subsequent group (N: < 50)]*

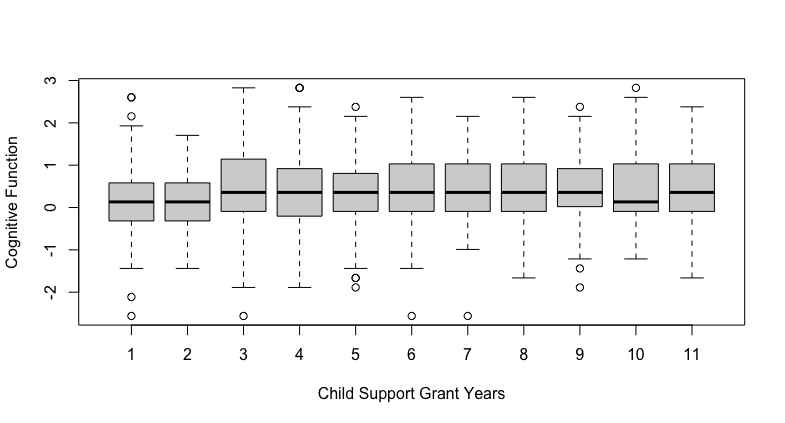

**S3 Fig: Cognitive function z scores are similar for those with 1 to 5 and 5.5 to 10 years of CSG eligibility while those with greater than 10 years of eligibility had relatively higher estimates for cognitive function**

*Step 4: Observing the similar estimates (from Fig S3) for cognitive function for 1 to 5 and 5.5 to 10 years of CSG eligibility and the relatively higher estimates for cognitive function for 10.5 years and above, we decided to place the cut-off at the 10-year mark, with “low” duration of CSG eligibility being ≤10 years and “high” duration of CSG eligibility being >10 years.*

**B. Sensitivity Analysis**

We analyzed CSG as a continuous variable (S1 Table) and dichotomized the years of CSG eligibility using two different thresholds, one at 8 years (S2 Table) and another at 12 years (S3 Table).

**S1 Table: Estimated effects of Child Support Grant eligibility (in years) on cognitive function in Child Support Grant-eligible mothers, Health and Ageing in Africa: A Longitudinal Study of an INDEPTH Community in South Africa, 2014/15**

| **Analytic Sample** | **Beta Coefficient (β)** | **95% Confidence Intervals** | **P value** |
| --- | --- | --- | --- |
| ***Full Sample (n= 944)*** | | |  |
| Unadjusted | 0.002 | -0.0005, 0.006 | 0.11 |
| Adjusted ^a^ | 0.0007 | -0.003, 0.005 | 0.71 |
| ***Lifetime number of children 1 to 4 (n = 390)*** | | |  |
| Unadjusted | 0.007 | -0.001, 0.01 | 0.07 |
| Adjusted ^b^ | 0.003 | -0.002, 0.009 | 0.17 |
| ***Lifetime number of children 5 and above (n = 554)*** | | | |
| Unadjusted | 0.004 | 0.0003, 0.007 | 0.03 |
| Adjusted ^b^ | -0.0008 | -0.006, 0.004 | 0.75 |

^a^ Adjusted for mother’s age and age squared, education (none, some primary, some secondary and more) and lifetime number of children (indicator variables)

^b^ Adjusted for mother’s age and age squared, education (none, some primary, some secondary and more) and lifetime number of children (indicator variables), within each lifetime children subgroup

**S2 Table: Estimated effects of high Child Support Grant eligibility (>8 years) compared to low Child Support Grant eligibility (≤8 years) on cognitive function in Child Support Grant-eligible mothers, Health and Ageing in Africa: A Longitudinal Study of an INDEPTH Community in South Africa, 2014/15**

| **Analytic Sample** | **Beta Coefficient (β)** | **95% Confidence Intervals** | **P value** |
| --- | --- | --- | --- |
| ***Full Sample (n= 944)*** | | |  |
| Unadjusted | 0.27 | 0.09, 0.44 | 0.003 |
| Adjusted ^a^ | 0.13 | -0.02, 0.28 | 0.08 |
| ***Lifetime number of children 1 to 4 (n = 390)*** | | |  |
| Unadjusted | 0.45 | 0.17, 0.73 | 0.001 |
| Adjusted ^b^ | 0.27 | 0.14, 0.39 | 0.007 |
| ***Lifetime number of children 5 and above (n = 554)*** | | | |
| Unadjusted | 0.13 | -0.03, 0.41 | 0.27 |
| Adjusted ^b^ | 0.03 | -0.21, 0.26 | 0.82 |

^a^ Adjusted for mother’s age and age squared, education (none, some primary, some secondary and more) and lifetime number of children (indicator variables)

^b^ Adjusted for mother’s age and age squared, education (none, some primary, some secondary and more) and lifetime number of children (indicator variables), within each lifetime children subgroup

**S3 Table: Estimated effects of high Child Support Grant eligibility (>12 years) compared to low Child Support Grant eligibility (≤12 years) on cognitive function in Child Support Grant-eligible mothers, Health and Ageing in Africa: A Longitudinal Study of an INDEPTH Community in South Africa, 2014/15**

| **Analytic Sample** | **Beta Coefficient (β)** | **95% Confidence Intervals** | **P value** |
| --- | --- | --- | --- |
| ***Full Sample (n= 944)*** | | |  |
| Unadjusted | 0.20 | 0.05, 0.35 | 0.009 |
| Adjusted ^a^ | 0.09 | -0.02, 0.19 | 0.09 |
| ***Lifetime number of children 1 to 4 (n = 390)*** | | |  |
| Unadjusted | 0.19 | -0.04, 0.43 | 0.11 |
| Adjusted ^b^ | 0.05 | -0.09, 0.19 | 0.33 |
| ***Lifetime number of children 5 and above (n = 554)*** | | | |
| Unadjusted | 0.23 | 0.04, 0.43 | 0.02 |
| Adjusted ^b^ | 0.11 | -0.06, 0.28 | 0.17 |

^a^ Adjusted for mother’s age and age squared, education (none, some primary, some secondary and more) and lifetime number of children (indicator variables)

^b^ Adjusted for mother’s age and age squared, education (none, some primary, some secondary and more) and lifetime number of children (indicator variables), within each lifetime children subgroup
