## Supplementary file2 for "Child Support Grant expansion and cognitive function among women in rural South Africa: findings from a natural experiment in HAALSI cohort"

| | Expansions | | <7[1] | | | | | | |
|---|---|---|---|---|---|---|---|---|---|
| | | **Jan-98** | **Apr-98** | **Jan-99** | **Apr-99** | **Jan-00** | **Apr-00** | **Jan-01** | **Apr-01** |
| | **Jan-89** | 9 | 9 | 10 | 10 | 11 | 11 | 12 | 12 |
| | **Jan-90** | 8 | 8 | 9 | 9 | 10 | 10 | 11 | 11 |
| | **Jan-91** | 7 | 7 | 8 | 8 | 9 | 9 | 10 | 10 |
| | **Jan-92** | 6 | 6 | 7 | 7 | 8 | 8 | 9 | 9 |
| | **Jan-93** | 5 | 5 | 6 | 6 | 7 | 7 | 8 | 8 |
| | **Jan-94** | 4 | 4 | 5 | 5 | 6 | 6 | 7 | 7 |
| | **Jan-95** | 3 | 3 | 4 | 4 | 5 | 5 | 6 | 6 |
| | **Jan-96** | 2 | 2 | 3 | 3 | 4 | 4 | 5 | 5 |
| | **Jan-97** | 1 | 1 | 2 | 2 | 3 | 3 | 4 | 4 |
| | **Jan-98** | 0 | 0 | 1 | 1 | 2 | 2 | 3 | 3 |
| | **Jan-99** | | | 0 | 0 | 1 | 1 | 2 | 2 |
| Birthyears | **Jan-00** | | | | | 0 | 0 | 1 | 1 |
| | **Jan-01** | | | | | | | 0 | 0 |
| | **Jan-02** | | | | | | | | |
| | **Jan-03** | | | | | | | | |
| | **Jan-04** | | | | | | | | |
| | **Jan-05** | | | | | | | | |
| | **Jan-06** | | | | | | | | |
| | **Jan-07** | | | | | | | | |
| | **Jan-08** | | | | | | | | |
| | **Jan-09** | | | | | | | | |
| | **Jan-10** | | | | | | | | |
| | **Jan-11** | | | | | | | | |
| | **Jan-12** | | | | | | | | |
| | **Jan-13** | | | | | | | | |
| | **Jan-14** | | | | | | | | |
| | **Jan-15** | | | | | | | | |
| | **Jan-16** | | | | | | | | |

| | | | <9[2] | | <11[2] | | <14[2] | | | | |
|---|---|---|---|---|---|---|---|---|---|---|---|
| **Jan-02** | **Apr-02** | **Jan-03** | **Apr-03** | **Jan-04** | **Apr-04** | **Jan-05** | **Apr-05** | **Jan-06** | **Apr-06** | **Jan-07** | **Apr-07** |
| 13 | 13 | 14 | 14 | 15 | 15 | 16 | 16 | 17 | 17 | 18 | 18 |
| 12 | 12 | 13 | 13 | 14 | 14 | 15 | 15 | 16 | 16 | 17 | 17 |
| 11 | 11 | 12 | 12 | 13 | 13 | 14 | 14 | 15 | 15 | 16 | 16 |
| 10 | 10 | 11 | 11 | 12 | 12 | 13 | 13 | 14 | 14 | 15 | 15 |
| 9 | 9 | 10 | 10 | 11 | 11 | 12 | 12 | 13 | 13 | 14 | 14 |
| 8 | 8 | 9 | 9 | 10 | 10 | 11 | 11 | 12 | 12 | 13 | 13 |
| 7 | 7 | 8 | 8 | 9 | 9 | 10 | 10 | 11 | 11 | 12 | 12 |
| 6 | 6 | 7 | 7 | 8 | 8 | 9 | 9 | 10 | 10 | 11 | 11 |
| 5 | 5 | 6 | 6 | 7 | 7 | 8 | 8 | 9 | 9 | 10 | 10 |
| 4 | 4 | 5 | 5 | 6 | 6 | 7 | 7 | 8 | 8 | 9 | 9 |
| 3 | 3 | 4 | 4 | 5 | 5 | 6 | 6 | 7 | 7 | 8 | 8 |
| 2 | 2 | 3 | 3 | 4 | 4 | 5 | 5 | 6 | 6 | 7 | 7 |
| 1 | 1 | 2 | 2 | 3 | 3 | 4 | 4 | 5 | 5 | 6 | 6 |
| 0 | 0 | 1 | 1 | 2 | 2 | 3 | 3 | 4 | 4 | 5 | 5 |
| | | 0 | 0 | 1 | 1 | 2 | 2 | 3 | 3 | 4 | 4 |
| | | | | 0 | 0 | 1 | 1 | 2 | 2 | 3 | 3 |
| | | | | | | 0 | 0 | 1 | 1 | 2 | 2 |
| | | | | | | | | 0 | 0 | 1 | 1 |
| | | | | | | | | | | 0 | 0 |

| | | <15[3] | | ≤16[4] | | | ≤17[4] | | <18[4] | | |
|---|---|---|---|---|---|---|---|---|---|---|---|
| **Jan-08** | **Apr-08** | **Jan-09** | **Apr-09** | **Jan-10** | **Apr-10** | **Jan-11** | **Apr-11** | **12-Jan** | **12-Apr** | **13-Jan** | **13-Apr** |
| 19 | 19 | 20 | 20 | 21 | 21 | 22 | 22 | 23 | 23 | 24 | 24 |
| 18 | 18 | 19 | 19 | 20 | 20 | 21 | 21 | 22 | 22 | 23 | 23 |
| 17 | 17 | 18 | 18 | 19 | 19 | 20 | 20 | 21 | 21 | 22 | 22 |
| 16 | 16 | 17 | 17 | 18 | 18 | 19 | 19 | 20 | 20 | 21 | 21 |
| 15 | 15 | 16 | 16 | 17 | 17 | 18 | 18 | 19 | 19 | 20 | 20 |
| 14 | 14 | 15 | 15 | 16 | 16 | 17 | 17 | 18 | 18 | 19 | 19 |
| 13 | 13 | 14 | 14 | 15 | 15 | 16 | 16 | 17 | 17 | 18 | 18 |
| 12 | 12 | 13 | 13 | 14 | 14 | 15 | 15 | 16 | 16 | 17 | 17 |
| 11 | 11 | 12 | 12 | 13 | 13 | 14 | 14 | 15 | 15 | 16 | 16 |
| 10 | 10 | 11 | 11 | 12 | 12 | 13 | 13 | 14 | 14 | 15 | 15 |
| 9 | 9 | 10 | 10 | 11 | 11 | 12 | 12 | 13 | 13 | 14 | 14 |
| 8 | 8 | 9 | 9 | 10 | 10 | 11 | 11 | 12 | 12 | 13 | 13 |
| 7 | 7 | 8 | 8 | 9 | 9 | 10 | 10 | 11 | 11 | 12 | 12 |
| 6 | 6 | 7 | 7 | 8 | 8 | 9 | 9 | 10 | 10 | 11 | 11 |
| 5 | 5 | 6 | 6 | 7 | 7 | 8 | 8 | 9 | 9 | 10 | 10 |
| 4 | 4 | 5 | 5 | 6 | 6 | 7 | 7 | 8 | 8 | 9 | 9 |
| 3 | 3 | 4 | 4 | 5 | 5 | 6 | 6 | 7 | 7 | 8 | 8 |
| 2 | 2 | 3 | 3 | 4 | 4 | 5 | 5 | 6 | 6 | 7 | 7 |
| 1 | 1 | 2 | 2 | 3 | 3 | 4 | 4 | 5 | 5 | 6 | 6 |
| 0 | 0 | 1 | 1 | 2 | 2 | 3 | 3 | 4 | 4 | 5 | 5 |
| | | 0 | 0 | 1 | 1 | 2 | 2 | 3 | 3 | 4 | 4 |
| | | | | 0 | 0 | 1 | 1 | 2 | 2 | 3 | 3 |
| | | | | | | 0 | 0 | 1 | 1 | 2 | 2 |
| | | | | | | | | 0 | 0 | 1 | 1 |
| | | | | | | | | | | 0 | 0 |

| 14-Jan | 14-Apr | 15-Jan | 15-Apr | 16-Jan | Yrs of eligibility | birthyears |
|---|---|---|---|---|---|---|
| 25 | 25 | 26 | 26 | 27 | 0 | **Jan-89** |
| 24 | 24 | 25 | 25 | 26 | 0 | **Jan-90** |
| 23 | 23 | 24 | 24 | 25 | 0 | **Jan-91** |
| 22 | 22 | 23 | 23 | 24 | 1 | **Jan-92** |
| 21 | 21 | 22 | 22 | 23 | 3 | **Jan-93** |
| 20 | 20 | 21 | 21 | 22 | 7.5 | **Jan-94** |
| 19 | 19 | 20 | 20 | 21 | 12.5 | **Jan-95** |
| 18 | 18 | 19 | 19 | 20 | 15 | **Jan-96** |
| 17 | 17 | 18 | 18 | 19 | 16.5 | **Jan-97** |
| 16 | 16 | 17 | 17 | 18 | 17.5 | **Jan-98** |
| 15 | 15 | 16 | 16 | 17 | 18 | **Jan-99** |
| 14 | 14 | 15 | 15 | 16 | 17 | **Jan-00** |
| 13 | 13 | 14 | 14 | 15 | 16 | **Jan-01** |
| 12 | 12 | 13 | 13 | 14 | 15 | **Jan-02** |
| 11 | 11 | 12 | 12 | 13 | 14 | **Jan-03** |
| 10 | 10 | 11 | 11 | 12 | 13 | **Jan-04** |
| 9 | 9 | 10 | 10 | 11 | 12 | **Jan-05** |
| 8 | 8 | 9 | 9 | 10 | 11 | **Jan-06** |
| 7 | 7 | 8 | 8 | 9 | 10 | **Jan-07** |
| 6 | 6 | 7 | 7 | 8 | 9 | **Jan-08** |
| 5 | 5 | 6 | 6 | 7 | 8 | **Jan-09** |
| 4 | 4 | 5 | 5 | 6 | 7 | **Jan-10** |
| 3 | 3 | 4 | 4 | 5 | 6 | **Jan-11** |
| 2 | 2 | 3 | 3 | 4 | 5 | **Jan-12** |
| 1 | 1 | 2 | 2 | 3 | 4 | **Jan-13** |
| 0 | 0 | 1 | 1 | 2 | 3 | **Jan-14** |
|  |  | 0 | 0 | 1 | 2 | **Jan-15** |
|  |  |  |  | 0 | 1 | **Jan-16** |

**References:**
1- The Evolution and Impact of Unconditional Cash Transfers in South Africa, Woolard and Leibbrandt, 201[illegible]
2 - The South African Child Support Grant Impact Assessment, 2012
3- Review of the Child Support Grant: Uses, Implementation and Obstacles, 2008
4 - Child support grant evaluation 2010, 2011

Expansions <7[1]

| | Apr-98 | Jan-99 | Apr-99 | Jan-00 | Apr-00 | Jan-01 | Apr-01 | Jan-02 |
|---|---|---|---|---|---|---|---|---|
| **Apr-89** | 9 | 9 | 10 | 10 | 11 | 11 | 12 | 12 |
| **Apr-90** | 8 | 8 | 9 | 9 | 10 | 10 | 11 | 11 |
| **Apr-91** | 7 | 7 | 8 | 8 | 9 | 9 | 10 | 10 |
| **Apr-92** | 6 | 6 | 7 | 7 | 8 | 8 | 9 | 9 |
| **Apr-93** | 5 | 5 | 6 | 6 | 7 | 7 | 8 | 8 |
| **Apr-94** | 4 | 4 | 5 | 5 | 6 | 6 | 7 | 7 |
| **Apr-95** | 3 | 3 | 4 | 4 | 5 | 5 | 6 | 6 |
| **Apr-96** | 2 | 2 | 3 | 3 | 4 | 4 | 5 | 5 |
| **Apr-97** | 1 | 1 | 2 | 2 | 3 | 3 | 4 | 4 |
| **Apr-98** | 0 | 0 | 1 | 1 | 2 | 2 | 3 | 3 |
| **Apr-99** | | | 0 | 0 | 1 | 1 | 2 | 2 |
| **Apr-00** | | | | | 0 | 0 | 1 | 1 |
| **Apr-01** | | | | | | | 0 | 0 |
| **Apr-02** | | | | | | | | |
| **Apr-03** | | | | | | | | |
| **Apr-04** | | | | | | | | |
| **Apr-05** | | | | | | | | |
| **Apr-06** | | | | | | | | |
| **Apr-07** | | | | | | | | |
| **Apr-08** | | | | | | | | |
| **Apr-09** | | | | | | | | |
| **Apr-10** | | | | | | | | |
| **Apr-11** | | | | | | | | |
| **Apr-12** | | | | | | | | |
| **Apr-13** | | | | | | | | |
| **Apr-14** | | | | | | | | |
| **Apr-15** | | | | | | | | |
| **Apr-16** | | | | | | | | |

| | | <9[2] | | <11[2] | | <14[2] | | |
|---|---|---|---|---|---|---|---|---|
| **Apr-02** | **Jan-03** | **Apr-03** | **Jan-04** | **Apr-04** | **Jan-05** | **Apr-05** | **Jan-06** | **Apr-06** |
| 13 | 13 | 14 | 14 | 15 | 15 | 16 | 16 | 17 |
| 12 | 12 | 13 | 13 | 14 | 14 | 15 | 15 | 16 |
| 11 | 11 | 12 | 12 | 13 | 13 | 14 | 14 | 15 |
| 10 | 10 | 11 | 11 | 12 | 12 | 13 | 13 | 14 |
| 9 | 9 | 10 | 10 | 11 | 11 | 12 | 12 | 13 |
| 8 | 8 | 9 | 9 | 10 | 10 | 11 | 11 | 12 |
| 7 | 7 | 8 | 8 | 9 | 9 | 10 | 10 | 11 |
| 6 | 6 | 7 | 7 | 8 | 8 | 9 | 9 | 10 |
| 5 | 5 | 6 | 6 | 7 | 7 | 8 | 8 | 9 |
| 4 | 4 | 5 | 5 | 6 | 6 | 7 | 7 | 8 |
| 3 | 3 | 4 | 4 | 5 | 5 | 6 | 6 | 7 |
| 2 | 2 | 3 | 3 | 4 | 4 | 5 | 5 | 6 |
| 1 | 1 | 2 | 2 | 3 | 3 | 4 | 4 | 5 |
| 0 | 0 | 1 | 1 | 2 | 2 | 3 | 3 | 4 |
| | | 0 | 0 | 1 | 1 | 2 | 2 | 3 |
| | | | | 0 | 0 | 1 | 1 | 2 |
| | | | | | | 0 | 0 | 1 |
| | | | | | | | | 0 |

| | | | | <15[3] | | ≤16[4] | | | ≤17[4] |
|---|---|---|---|---|---|---|---|---|---|
| **Jan-07** | **Apr-07** | **Jan-08** | **Apr-08** | **Jan-09** | **Apr-09** | **Jan-10** | **Apr-10** | **Jan-11** | **Apr-11** |
| 17 | 18 | 18 | 19 | 19 | 20 | 20 | 21 | 21 | 22 |
| 16 | 17 | 17 | 18 | 18 | 19 | 19 | 20 | 20 | 21 |
| 15 | 16 | 16 | 17 | 17 | 18 | 18 | 19 | 19 | 20 |
| 14 | 15 | 15 | 16 | 16 | 17 | 17 | 18 | 18 | 19 |
| 13 | 14 | 14 | 15 | 15 | 16 | 16 | 17 | 17 | 18 |
| 12 | 13 | 13 | 14 | 14 | 15 | 15 | 16 | 16 | 17 |
| 11 | 12 | 12 | 13 | 13 | 14 | 14 | 15 | 15 | 16 |
| 10 | 11 | 11 | 12 | 12 | 13 | 13 | 14 | 14 | 15 |
| 9 | 10 | 10 | 11 | 11 | 12 | 12 | 13 | 13 | 14 |
| 8 | 9 | 9 | 10 | 10 | 11 | 11 | 12 | 12 | 13 |
| 7 | 8 | 8 | 9 | 9 | 10 | 10 | 11 | 11 | 12 |
| 6 | 7 | 7 | 8 | 8 | 9 | 9 | 10 | 10 | 11 |
| 5 | 6 | 6 | 7 | 7 | 8 | 8 | 9 | 9 | 10 |
| 4 | 5 | 5 | 6 | 6 | 7 | 7 | 8 | 8 | 9 |
| 3 | 4 | 4 | 5 | 5 | 6 | 6 | 7 | 7 | 8 |
| 2 | 3 | 3 | 4 | 4 | 5 | 5 | 6 | 6 | 7 |
| 1 | 2 | 2 | 3 | 3 | 4 | 4 | 5 | 5 | 6 |
| 0 | 1 | 1 | 2 | 2 | 3 | 3 | 4 | 4 | 5 |
| | 0 | 0 | 1 | 1 | 2 | 2 | 3 | 3 | 4 |
| | | | 0 | 0 | 1 | 1 | 2 | 2 | 3 |
| | | | | | 0 | 0 | 1 | 1 | 2 |
| | | | | | | | 0 | 0 | 1 |
| | | | | | | | | | 0 |

| | <18[4] | | | | | | | | |
|---|---|---|---|---|---|---|---|---|---|
| 12-Jan | 12-Apr | 13-Jan | 13-Apr | 14-Jan | 14-Apr | 15-Jan | 15-Apr | 16-Jan | Yrs of eligibi |
| 22 | 23 | 23 | 24 | 24 | 25 | 25 | 26 | 26 | 0 |
| 21 | 22 | 22 | 23 | 23 | 24 | 24 | 25 | 25 | 0 |
| 20 | 21 | 21 | 22 | 22 | 23 | 23 | 24 | 24 | 0 |
| 19 | 20 | 20 | 21 | 21 | 22 | 22 | 23 | 23 | 2 |
| 18 | 19 | 19 | 20 | 20 | 21 | 21 | 22 | 22 | 4.5 |
| 17 | 18 | 18 | 19 | 19 | 20 | 20 | 21 | 21 | 10 |
| 16 | 17 | 17 | 18 | 18 | 19 | 19 | 20 | 20 | 14 |
| 15 | 16 | 16 | 17 | 17 | 18 | 18 | 19 | 19 | 16 |
| 14 | 15 | 15 | 16 | 16 | 17 | 17 | 18 | 18 | 17 |
| 13 | 14 | 14 | 15 | 15 | 16 | 16 | 17 | 17 | 18 |
| 12 | 13 | 13 | 14 | 14 | 15 | 15 | 16 | 16 | 17.5 |
| 11 | 12 | 12 | 13 | 13 | 14 | 14 | 15 | 15 | 16.5 |
| 10 | 11 | 11 | 12 | 12 | 13 | 13 | 14 | 14 | 15.5 |
| 9 | 10 | 10 | 11 | 11 | 12 | 12 | 13 | 13 | 14.5 |
| 8 | 9 | 9 | 10 | 10 | 11 | 11 | 12 | 12 | 13.5 |
| 7 | 8 | 8 | 9 | 9 | 10 | 10 | 11 | 11 | 12.5 |
| 6 | 7 | 7 | 8 | 8 | 9 | 9 | 10 | 10 | 11.5 |
| 5 | 6 | 6 | 7 | 7 | 8 | 8 | 9 | 9 | 10.5 |
| 4 | 5 | 5 | 6 | 6 | 7 | 7 | 8 | 8 | 9.5 |
| 3 | 4 | 4 | 5 | 5 | 6 | 6 | 7 | 7 | 8.5 |
| 2 | 3 | 3 | 4 | 4 | 5 | 5 | 6 | 6 | 7.5 |
| 1 | 2 | 2 | 3 | 3 | 4 | 4 | 5 | 5 | 6.5 |
| 0 | 1 | 1 | 2 | 2 | 3 | 3 | 4 | 4 | 5.5 |
| | 0 | 0 | 1 | 1 | 2 | 2 | 3 | 3 | 4.5 |
| | | | 0 | 0 | 1 | 1 | 2 | 2 | 3.5 |
| | | | | | 0 | 0 | 1 | 1 | 2.5 |
| | | | | | | | 0 | 0 | 1.5 |
| | | | | | | | | | 0.5 |

| birthyears |
| --- |
| **Apr-89** |
| **Apr-90** |
| **Apr-91** |
| **Apr-92** |
| **Apr-93** |
| **Apr-94** |
| **Apr-95** |
| **Apr-96** |
| **Apr-97** |
| **Apr-98** |
| **Apr-99** |
| **Apr-00** |
| **Apr-01** |
| **Apr-02** |
| **Apr-03** |
| **Apr-04** |
| **Apr-05** |
| **Apr-06** |
| **Apr-07** |
| **Apr-08** |
| **Apr-09** |
| **Apr-10** |
| **Apr-11** |
| **Apr-12** |
| **Apr-13** |
| **Apr-14** |
| **Apr-15** |
| **Apr-16** |

**References:**

1- The Evolution and Impact of Unconditional Cash Transfers in South Africa, Woolard and Leibbrandt, 201[illegible]
2 - The South African Child Support Grant Impact Assessment, 2012
3- Review of the Child Support Grant: Uses, Implementation and Obstacles, 2008
4 - Child support grant evaluation 2010, 2011
